# The Movember Global Action Plan 1 (GAP1) - Unique Prostate Cancer Tissue Microarray Resource

**DOI:** 10.1101/2021.06.03.21256653

**Authors:** Véronique Ouellet, Andrew Erickson, Kathy Wiley, Colm Morrissey, Viktor Berge, Carlos S. Moreno, Kristin Austlid Tasken, Dominique Trudel, Lawrence D. True, Michael S. Lewis, Aud Svindland, Onur Ertunc, Igor Damasceno Vidal, Adeboye O. Osunkoya, Tracy Jones, G. Steven Bova, Tarja Lamminen, Ariel H. Achtman, Mark Buzza, Michelle M Kouspou, GAP1 UTMAs Contributing Investigators, Steven A Bigler, Xinchun Zhou, Stephen J. Freedland, Anne-Marie Mes-Masson, Isla P. Garraway, Bruce J. Trock, Pekka Taimen, Fred Saad, Tuomas Mirtti, Beatrice S. Knudsen, Angelo M. De Marzo

## Abstract

**Background:** The need to better understand the molecular underpinnings of the heterogeneous outcomes of patients with prostate cancer is a pressing global problem and a key research priority for Movember. To address this, the Movember Global Action Plan 1 Unique tissue microarray (GAP1-UTMA) project constructed a set of unique and richly annotated TMAs from prostate cancer samples obtained from multiple institutions across several global locations.

**Methods:** Three separate TMA sets were built that differ by purpose and disease state.

**Results:** The intended use of TMA1 is to validate biomarkers that help determine which clinically localized prostate cancers with associated lymph node metastasis have a high risk of progression to lethal castration resistant metastatic disease, and to compare molecular properties of high risk index lesions within the prostate to regional lymph node metastases resected at the time of prostatectomy. TMA2 was designed to address questions regarding risk of castration resistant prostate cancer (CRPC) and response to suppression of the androgen receptor/androgen axis, and characterization of the castration-resistant phenotype. TMA3’s intended use is to assess and better understand the heterogeneity of molecular markers across different anatomic sites in lethal prostate cancer metastases.

**Conclusion:** The GAP1-UTMA project has succeeded in combining a large set of rare tissue specimens from 501 prostate cancer patients with rich clinical annotation.

**Impact:** This resource is now available to the prostate cancer community as a tool for biomarker validation to address important unanswered clinical questions around disease progression and response to treatment.

## Introduction

Prostate cancer is the second most common cancer and the sixth leading cause of cancer death worldwide in men (1,2). While some prostate cancers remain clinically indolent for many years, others progress at a variable rate to lethal metastatic castration resistant disease and death. The death rate from prostate cancer has decreased in the last two decades, potentially as a result of better detection and treatment of clinically localized disease. However, despite this progress many men with tumors of low inherent aggressiveness are over-treated, while others with intermediate risk or high risk localized disease are undertreated (3–5). The mainstay for clinical decisions for localized prostate cancer is a biopsy followed by Gleason grading of the tumor. Gleason grading remains a powerful predictor of outcome in the extremes and those patients with Gleason Grade Group 1 (Gleason score 6) and some with low volume Grade Group 2 (3+4=7) are forgoing immediate treatment and increasingly enrolling in active surveillance programs. However, for those patients with clinically localized higher risk disease (high volume cancers of Grade Group 2 and higher, and any patients with Grade Groups 4-5), a significant fraction develop disease recurrence and progression after attempts at definitive local treatments(6). For these patients, additional treatments are needed, yet there are no standard adjuvant therapies for localized prostate cancer. Further, for those patients with intermediate risk disease, the clinical course is quite variable and not well predicted by Gleason grading and clinical staging.

Presently, while a number of different molecular biomarkers have shown promise (7–9), there are no routinely employed validated biomarkers to differentiate indolent from aggressive intermediate risk cases that can be used to enhance decision making. A subgroup of patients with clinically localized disease have prostate cancer which has spread to regional lymph nodes, which is often discovered incidentally when these lymph nodes are removed during a radical prostatectomy (RP) and examined microscopically. Interestingly, the presence of regional lymph node metastasis does not always portend an aggressive clinical course because some patients in this disease state remain stable without overt new metastatic disease and development of bone or soft tissue metastases for many years (10,11). Indeed, some patients who are treated with Androgen Deprivation Therapy (ADT), for either localized or metastatic disease, can have highly prolonged responses, while others succumb to castration-resistant prostate cancer (CRPC) quite quickly (12). Lastly, when untreated, only a third of patients with biochemical recurrence (BCR) will develop clinically significant metastatic disease in a 7-year follow-up period (13), which underlines the relatively poor performance of BCR as a surrogate marker for prostate cancer survival outcomes. More precise biomarkers are needed to determine which of these locally treated cancers are likely to progress and/or develop resistance to ADT.

Another clinical problem that occurs in late stage disease, often after several systemic therapies involving androgen signaling deprivation and/or chemotherapy, is disease heterogeneity(14–16) with some lesions responding to specific systemic agents and others not. This heterogeneity in CRPC is similar to many other types of late stage metastatic cancer and improvements in deciphering its molecular features and mechanisms require tissue sampling from multiple metastatic sites, which is generally quite difficult outside of autopsies. The development and validation of biomarkers that can help better stratify risk in men with localized disease, to better understand who may rapidly progress on ADT, and to enhance our knowledge regarding the molecular features of late stage disease heterogeneity was the main rationale for initiating this Movember Global Action Plan 1 (GAP1) unique tissue microarray (TMA) project (GAP1-UTMA).

While TMAs have been used in more than 2000 publications in prostate cancer, the majority of these studies have been carried out by individual organizations/investigators and focused on the prognostic value of biomarkers, often using rising serum PSA as an indicator of poor outcome. Movember’s Global Action Plans (GAP), launched in 2011 with the GAP1 biomarker project, were established to address critical challenges in prostate cancer research through global collaboration. Movember identified a need to create collections of valuable tissue resources with clinical annotation to support the first batch of GAP1 projects to help improve our understanding of the biology of treatment response and resistance and validating promising prostate cancer tissue-based biomarkers. In this GAP1 collaborative initiative, we assembled an international team with multidisciplinary expertise to pool diverse tissue sample resources to develop a novel set of TMAs consisting of prostate cancer tissue specimens from multiple disease states. This robust resource could not have been assembled using samples from any single institution.

## Materials and methods

### Patients and specimen collection

Formalin-fixed paraffin-embedded prostate cancer patient specimens (prostate, lymph nodes and other metastatic sites) used for TMA construction were retrieved from each participating hospital’s pathology department where patients underwent surgery, transurethral resection of the prostate (TURP) or biopsy between 1978-2016. These hospitals included the Centre hospitalier de l’Université de Montréal (CHUM), Johns Hopkins Hospital (JHU), Helsinki University Hospital (HUS), Institute of Biomedicine, University of Turku and Turku University Hospital (TYKS), Oslo University Hospital (OUH), University of Mississippi Baptist Medical Centre (UMBMC), Greater Los Angeles VA Healthcare System, Durham VA Health Care System, Emory University, and the University of Washington. For autopsy cases, all specimens were processed within 8 h (University of Washington), 14 h (JHU) or 24 h (Oslo University Hospital) of death. Specimens were fixed in buffered formalin and embedded in paraffin. Bone metastases were decalcified in 10% formic acid before embedding.

All centers received ethical review board permission to use patient material and conduct this study Comité d’éthique de la recherche du CHUM (CE.14.128), the Institutional Review Board of the : JHU School of Medicine’s, the UMBMC, the VA Greater Los Angeles (Department of Veterans Affairs PCC#2015-040408), the Ethics Committee of Hospital District of Helsinki and Uusimaa (84/13/03/00/2014; §3 30.01.2015), the Hospital District of Southwest Finland (number T206/2014) National Supervisory Authority for Welfare and Health for HUS and TYKS (VALVIRA, 8008/06.01.03/2014), the Regional Committees for Medical and Health Research Ethics for OUH (REC 2013/1713) and Center for Healthcare Ethics for Cedar Sinai (Pro00033387 and Pro00020577). Patient informed consents were obtained as required by individual institutional ethical review boards (CHUM CE12.216, HUS, TYKS and University of Washington, Cedar Sinai) or waivers were granted (JHU, OUH, both VA Health Care System and UMBMC).

### Histology review

The hematoxylin and eosin (H&E) or Weigert van Gieson stained sections of the whole prostates and pelvic lymph nodes from RP for TMA1, biopsies, RP and TURPs for TMA2 and prostates or metastatic lesions obtained at autopsy for TMA3 were reviewed by a pathologist within each institution. For the RP specimens, the pathologists identified the index tumor (largest tumor or the highest grade nodule). For other tissue types, the overall tissue quality and regions of high tumor content with minimal necrosis were selected for inclusion. Of note, some of the metastases from TMA3 have variably large regions of necrosis and some metastases have very few tumor cells, such as those from osteoblastic bones. Patient and tissue block selection were performed using specific guidelines.

### TMA Design, Construction and review

TMAs were constructed according to standard operating procedures (SOP) developed at each site and reviewed centrally to assure homogeneity prior to TMA construction. To build each TMA block, CHUM and Cedars-Sinai (Greater Los Angeles VA and Durham VA Health Care System, Emory University) used the TMArrayer (Pathology Devices, Inc., Westminster, MD, USA), while HUS and TYKS used Quick Ray Manual Tissue Microarrayer Full Set (Unitma, Seoul, Korea). The University of Oslo utilized a semi-automated Beecher Instrument; TMABooster OI (Alphelys, Plaisir, France). JHU utilized the Estigen Tissue Science Manual Tissue Arrayer MTA-1 (Tartu, Estonia; formerly Beecher instruments). The University of Washington utilized the Estigen Tissue Science Manual Tissue Arrayer MTA-1 (Tartu, Estonia; formerly Beecher instruments). The University of Mississippi Medical Center also used the Estigen Tissue Science Manual Tissue Arrayer MTA-1 (Tartu, Estonia; formerly Beecher instruments).

A total of three cores from prostatic tissue (0.6mm or 1mm), 2-3 cores from lymph nodes (0.6mm or 1mm) and other metastatic lesions (0.6 mm or 1mm) were transferred to a recipient TMA block in a serial manner (non-randomized). For biopsy tissue from prostate or metastatic lesions (0.6mm), at least 2 cores from each specimen were also arrayed in a serial manner (non-randomized) on a separate recipient TMA block. For each TMA category (TMA 1, 2 and 3), each institution constructed a Test TMA, that consisted of a subset of the same specimens used in the full TMAs.

For each TMA, we included control tissues provided by Cedars-Sinai Medical Center (CSMC) (human tonsil, kidney, colon and liver) and CHUM (xenografts containing 22rv1, PC3, LNCaP and DU145 prostate cancer cell lines injected into and grown in immunocompromised/nude mice). An institutional approval was provided for the use of the human control tissue specimens. Mouse xenograft experiments were performed according to institutional rules and following approbation of the protocol by the Comité institutionnel de protection des animaux. Each of the control tissues (mouse and human) was divided into eight pieces, fixed in formalin, paraffin embedded and sent to the individual institutions to include in each TMA block. This process of using centrally processed control cell lines and tissues provides a quality control measure of TMA slide staining to detect batch effects in staining due to tissue processing protocols (instead of underlying biology). Additionally, for many IHC stains there is a wealth of prior information regarding phenotypic features of these cell lines/xenografts and the common human tissues. To demarcate the starting X and Y coordinates (e.g. the upper left corner of the TMA block) control tissues (human kidney or liver, depending upon institutional preference) were arranged either outside of or just within the main X and Y coordinates to facilitate proper orientation of each TMA slide. Quality control of H&E stained sections from each TMA was performed by local and central genito-urinary pathologists.

### TMA Data Handling

Each patient was assigned a specific code, consisting of a combined unique deidentified Specimen ID and Institution-specific 3 letter ID. This information was shared with the coordinating center (JHU) and was used to enter the data into the TMAJ Database (TMAJ), an open-source software system designed to support TMA pathology data (17,18) (http://tmaj.pathology.jhmi.edu/). The TMAJ database tracks the specimen ID, and institution and information for each FFPE block (e.g. anatomic site of origin) that is used for TMA punching and provides an export of a TMA map that was used for all sites for quality control. Clinical data associated with specimens were collected in a spreadsheet containing predefined data elements (**Supplemental Table 1**) with specific definitions to promote harmonization of data between sites. No HIPAA-defined protected health information is included in the data. These data were sent to the central repository where they were reviewed for consistency and when inconsistencies were identified, they were rectified by communication with the initiating site and modified if needed. Revised clinical data from each site were collated into a unique SAS database. These data are stored on a secure server in the central repository and are linked to the specimen data in the TMAs using the same unique identifier (Specimen ID and institutional ID) mentioned above. The combined clinical data from all sites are only accessible to the study biostatistician and research coordinator at the Coordinating Center. Each TMA slide from each TMA block is assigned a unique ID in the TMAJ database, and data about which stain was performed, including the site, antibody and experimental details are recorded.

### Storage of TMA Blocks, Unstained Slides and Whole Slide Images

Each TMA block is kept at room temperature within its respective institution due to legal and ethical restrictions. TMA blocks were subjected to sectioning (N=20 slides) at each respective institution and the H&E section as well as the remaining 19 unstained slides of each TMA were sent to the coordinating center. Following reception, the H&E slides were subjected to whole slide scanning using a Hamamatsu Photonics NanoZoomer XR, SN 510076 instrument and whole slide scan image files were uploaded to a web-based slide image management system (Concentriq™ from PROSCIA) and shared with the other sites. The remaining unstained TMA slides are stored in a plastic bag in a monitored −20°C freezer according to PCBN program SOPs (18), until requested by approved investigators.

### Immunohistochemistry and Scoring

Each test TMA slide was stained for the following: recombinant anti-PSMA (Abcam, Cambridge, MA, clone EPR 6253, rabbit monoclonal, dilution 1:300), PTEN (Cell Signaling, Danvers, MA, clone D4.3 XP, rabbit monoclonal, dilution 1:100), anti-ERG (Roche, Indianapolis, IN, clone EPR3864, rabbit monoclonal, 23ug/ml), AR (androgen receptor, Cell Signaling, Danvers, MA, clone D6F11, rabbit monoclonal, dilution 1:400), PSA (Prostate Specific Antigen, Santa Clara, CA, clone ER-PR8, monoclonal anti-mouse, dilution 1:50) and NKX3.1 (rabbit polyclonal (19), dilution 1:1000). The slides were stained using the Ventana automated platform (Ventana Discovery Ultra HQ-HRP Hapten Detection, Ventana Medical System, Tucson, AZ). The stained TMA sections were scanned as above for H&E slides using the Hamamatsu NanoZoomer, stored on Consentriq™ (PROSCIA), and shared across all sites. For scoring the IHC staining, pathologists with expertise in prostate cancer reviewed the scanned slides and scored each spot. For every marker, staining was considered positive or negative. Staining was considered negative if there was a complete absence of staining in the tumor cells. Positive staining in tumor cells was categorized as homogeneous or heterogeneous, with heterogeneity meaning either a difference in the fraction of positive tumor cells, the intensity of the staining across the tumor cells or both. For scoring of PTEN and ERG, if there was a complete absence of staining in tumor cells and surrounding stromal cells, then that given TMA spot was considered inadequate for scoring and was not included. PTEN staining was considered ambiguous and was not scored when negative staining in the tumor was associated with only very weak positive, or negative staining in surrounding stromal cells. Similar methods for PTEN staining and scoring have been published previously(20,21). TMA data was recorded at each site by the study pathologist for each marker by using a google sheet with pull down menus for diagnoses and scoring. For each test-TMA stained slide the data was then consolidated into a larger excel spreadsheet for initial data tabulation in SAS and separately in STATA 15.

### Statistical analyses

In the present study we performed IHC staining for 6 different biomarkers in the test TMAs, primarily for quality control and proof of concept purposes. For each test TMA, we obtained descriptive summary statistics of the proportion of cases staining negatively, and positively (homogeneous and heterogeneous) and compared scoring across tissue types using Fisher’s exact test or chi-square. For example, for test TMA 1, univariate analyses were used to correlate biomarkers to each other. An example of this is the determination of the correlation between AR and its known downstream targets NKX3.1 and PSA, which gives a measure of AR transcriptional activity. For test TMAs 2 and 3, descriptive statistics are employed only. All statistical analyses were carried out using SAS v9.4 (SAS Institute, Cary, NC).

## Results

### Brief Descriptions of TMAs and Demographics

The main objective of the GAP1 Unique TMAs was to create a TMA-based resource to address three outstanding questions. The purpose of each TMAs and their intent of use as well as the details regarding the inclusion and exclusion criteria are presented in **Table 1a**. The TMA composition including number of TMA blocks, specimens, tissue type specification and tissue core numbers are described in **Table 1b**. Demographic and clinico-pathological data for each TMA series are presented in **Table 2**.

**Table 1:**
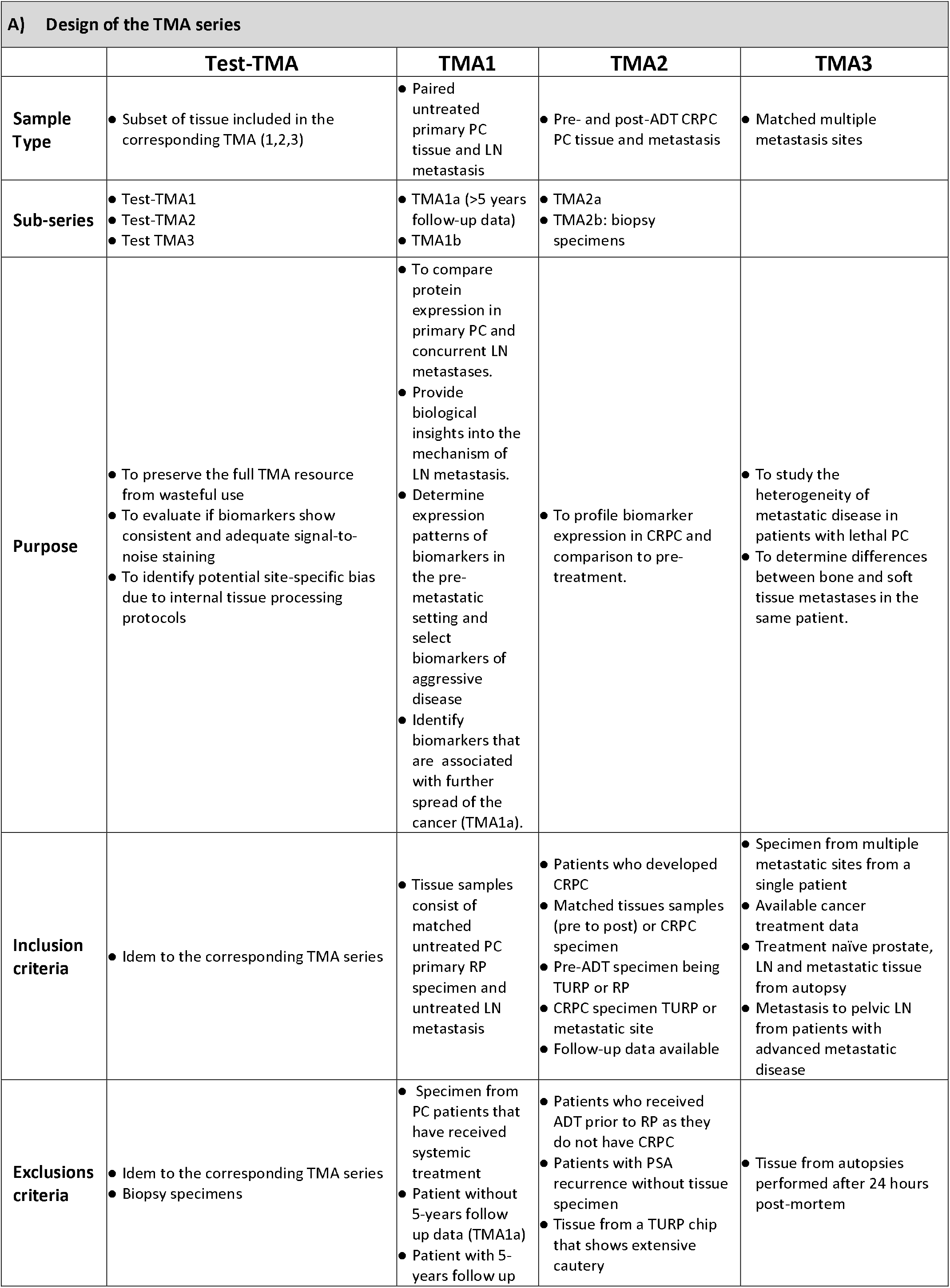
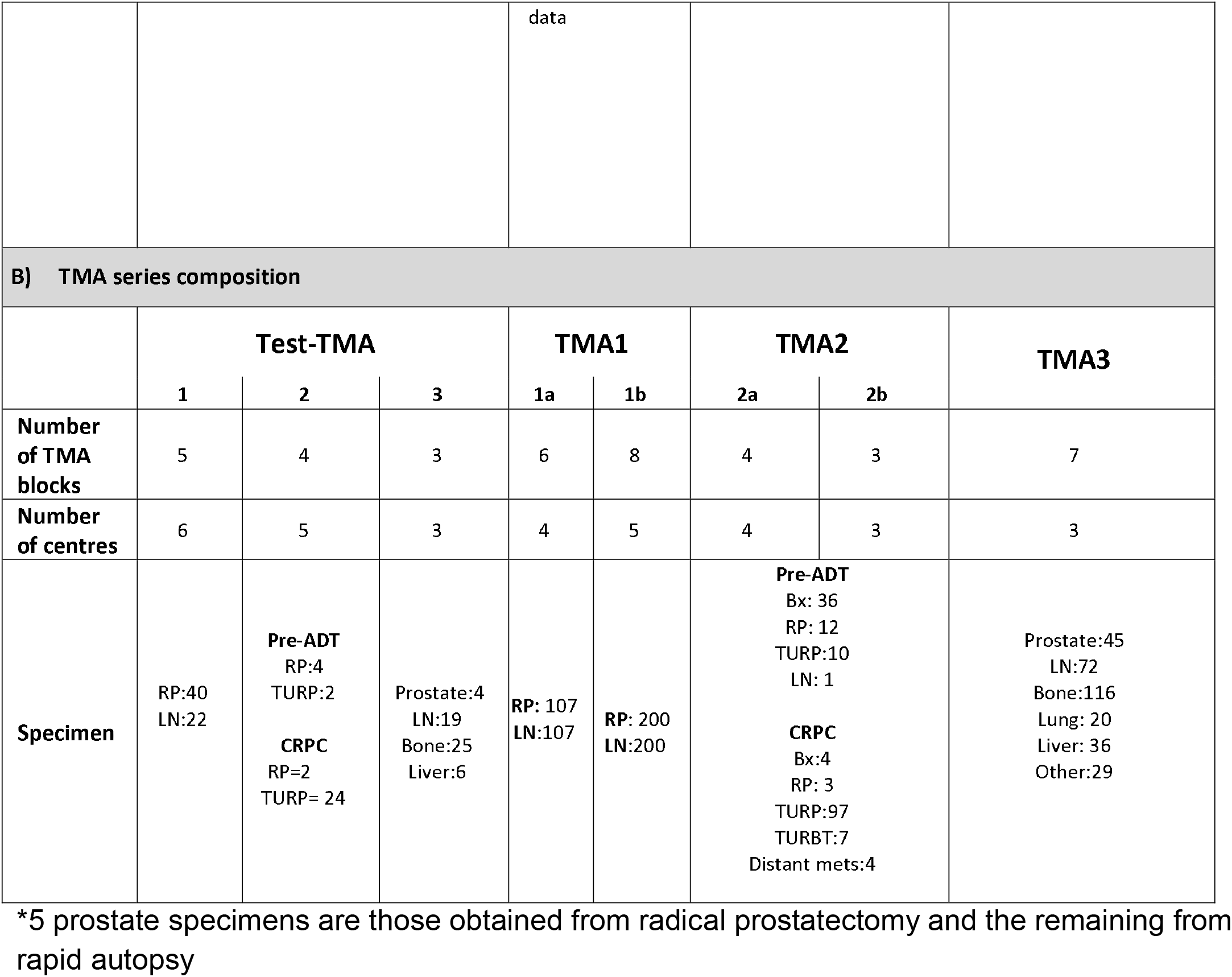
Unique TMA Description

| A) Design of the TMA series |  |  |  |  |
| --- | --- | --- | --- | --- |
|  | Test-TMA | TMA1 | TMA2 | TMA3 |
| **Sample Type** | Subset of tissue included in the corresponding TMA (1,2,3) | Paired untreated primary PC tissue and LN metastasis | Pre- and post-ADT CRPC PC tissue and metastasis | Matched multiple metastasis sites |
| **Sub-series** | Test-TMA1<br>Test-TMA2<br>Test TMA3 | TMA1a (>5 years follow-up data)<br>TMA1b | TMA2a<br>TMA2b: biopsy specimens |  |
| **Purpose** | To preserve the full TMA resource from wasteful use<br>To evaluate if biomarkers show consistent and adequate signal-to-noise staining<br>To identify potential site-specific bias due to internal tissue processing protocols | To compare protein expression in primary PC and concurrent LN metastases.<br>Provide biological insights into the mechanism of LN metastasis.<br>Determine expression patterns of biomarkers in the pre-metastatic setting and select biomarkers of aggressive disease<br>Identify biomarkers that are associated with further spread of the cancer (TMA1a). | To profile biomarker expression in CRPC and comparison to pre-treatment. | To study the heterogeneity of metastatic disease in patients with lethal PC<br>To determine differences between bone and soft tissue metastases in the same patient. |
| **Inclusion criteria** | Idem to the corresponding TMA series | Tissue samples consist of matched untreated PC primary RP specimen and untreated LN metastasis | Patients who developed CRPC<br>Matched tissues samples (pre to post) or CRPC specimen<br>Pre-ADT specimen being TURP or RP<br>CRPC specimen TURP or metastatic site<br>Follow-up data available | Specimen from multiple metastatic sites from a single patient<br>Available cancer treatment data<br>Treatment naïve prostate, LN and metastatic tissue from autopsy<br>Metastasis to pelvic LN from patients with advanced metastatic disease |
| **Exclusions criteria** | Idem to the corresponding TMA series<br>Biopsy specimens | Specimen from PC patients that have received systemic treatment<br>Patient without 5-years follow up data (TMA1a)<br>Patient with 5-years follow up | Patients who received ADT prior to RP as they do not have CRPC<br>Patients with PSA recurrence without tissue specimen<br>Tissue from a TURP chip that shows extensive cautery | Tissue from autopsies performed after 24 hours post-mortem |

|  |  |  |  |  |  |  |  |  |
| --- | --- | --- | --- | --- | --- | --- | --- | --- |
|  |  |  | data |  |  |  |  |  |
| B) TMA series composition |  |  |  |  |  |  |  |  |
|  | Test-TMA |  |  | TMA1 |  | TMA2 |  | TMA3 |
|  | 1 | 2 | 3 | 1a | 1b | 2a | 2b |  |
| Number of TMA blocks | 5 | 4 | 3 | 6 | 8 | 4 | 3 | 7 |
| Number of centres | 6 | 5 | 3 | 4 | 5 | 4 | 3 | 3 |
| Specimen | RP:40<br>LN:22 | Pre-ADT<br>RP:4<br>TURP:2<br>CRPC<br>RP=2<br>TURP= 24 | Prostate:4<br>LN:19<br>Bone:25<br>Liver:6 | RP: 107<br>LN:107 | RP: 200<br>LN:200 | Pre-ADT<br>Bx: 36<br>RP: 12<br>TURP:10<br>LN: 1<br>CRPC<br>Bx:4<br>RP: 3<br>TURP:97<br>TURBT:7<br>Distant mets:4 |  | Prostate:45<br>LN:72<br>Bone:116<br>Lung: 20<br>Liver: 36<br>Other:29 |

\*5 prostate specimens are those obtained from radical prostatectomy and the remaining from rapid autopsy

**Table 2:**
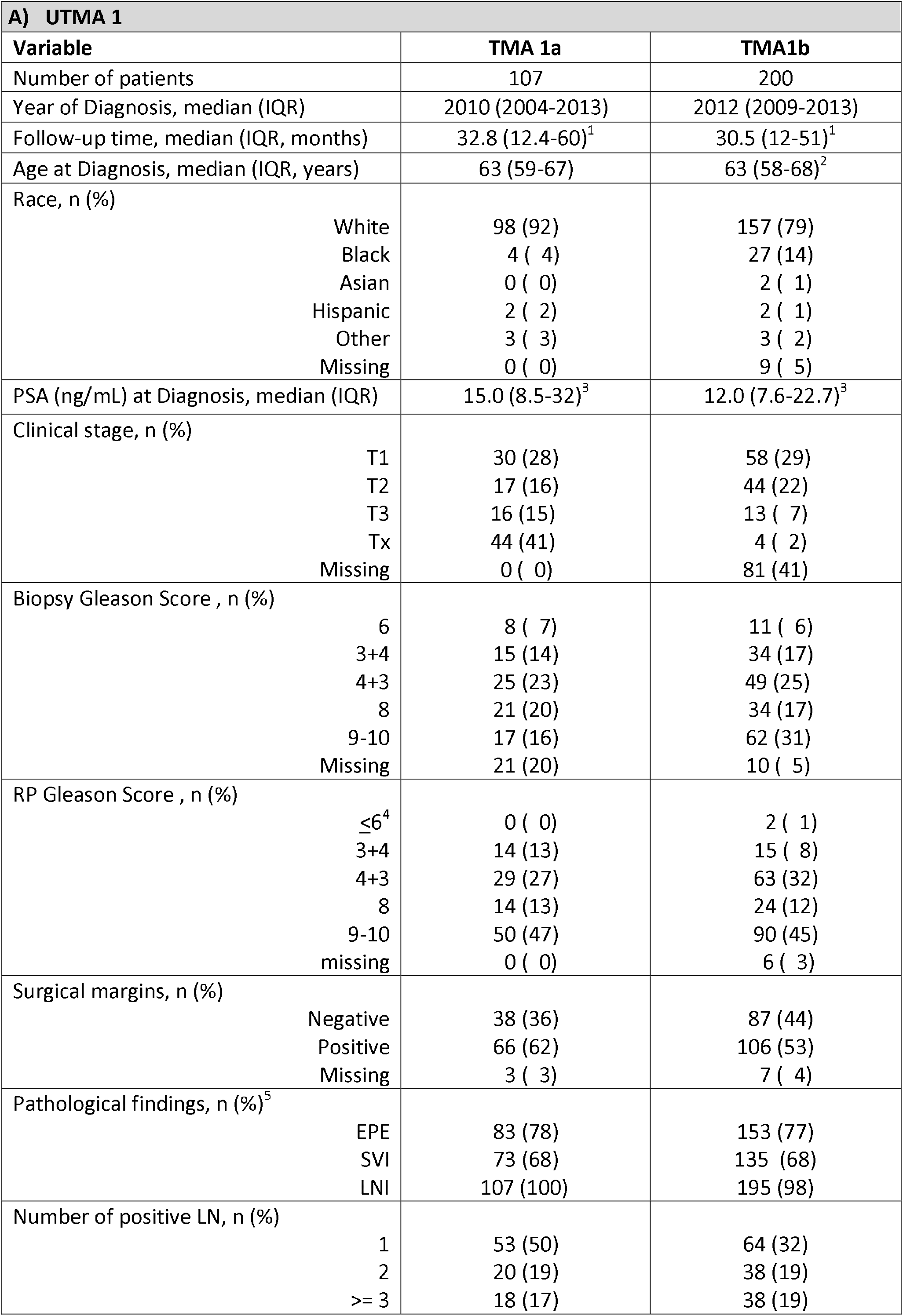
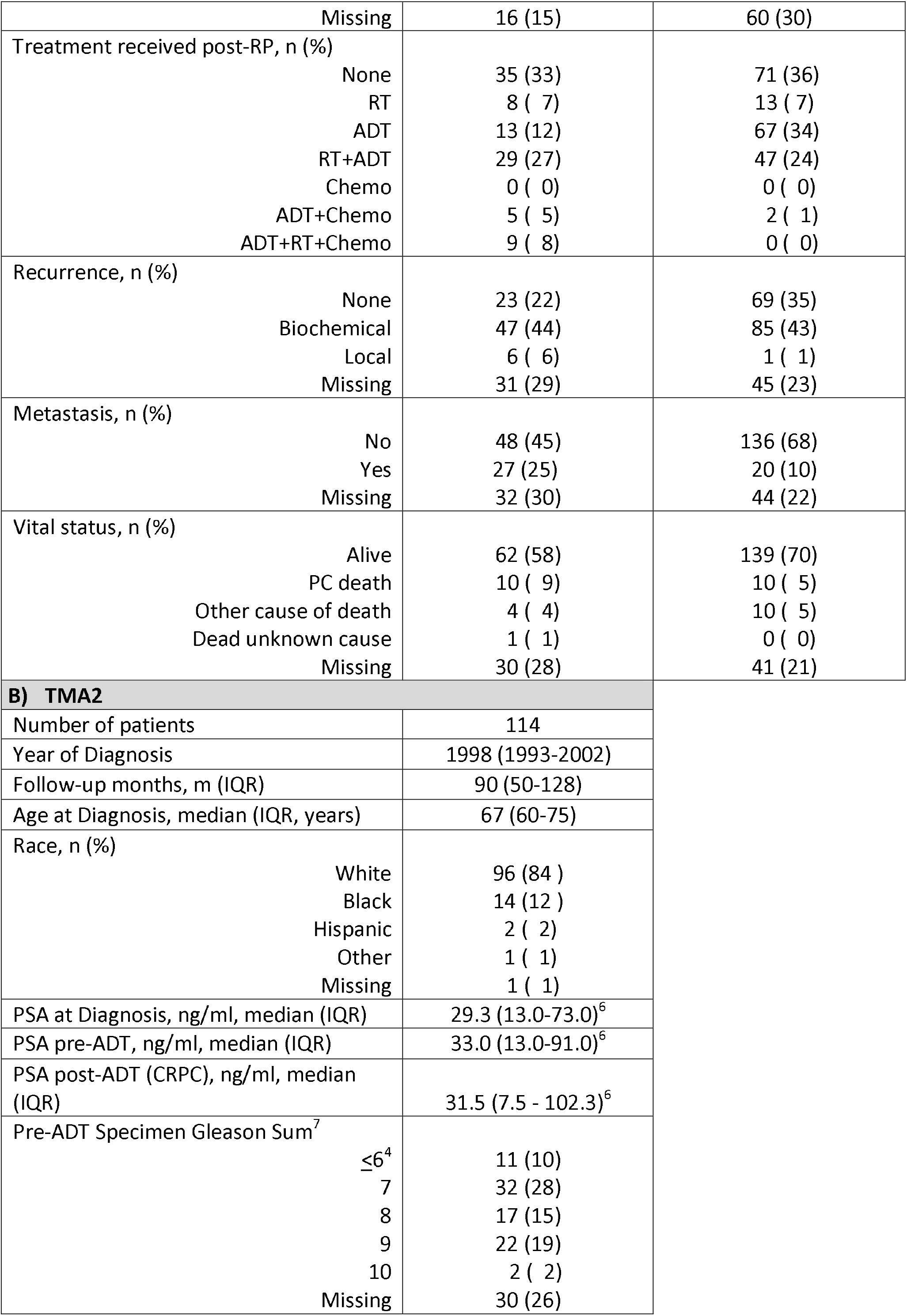
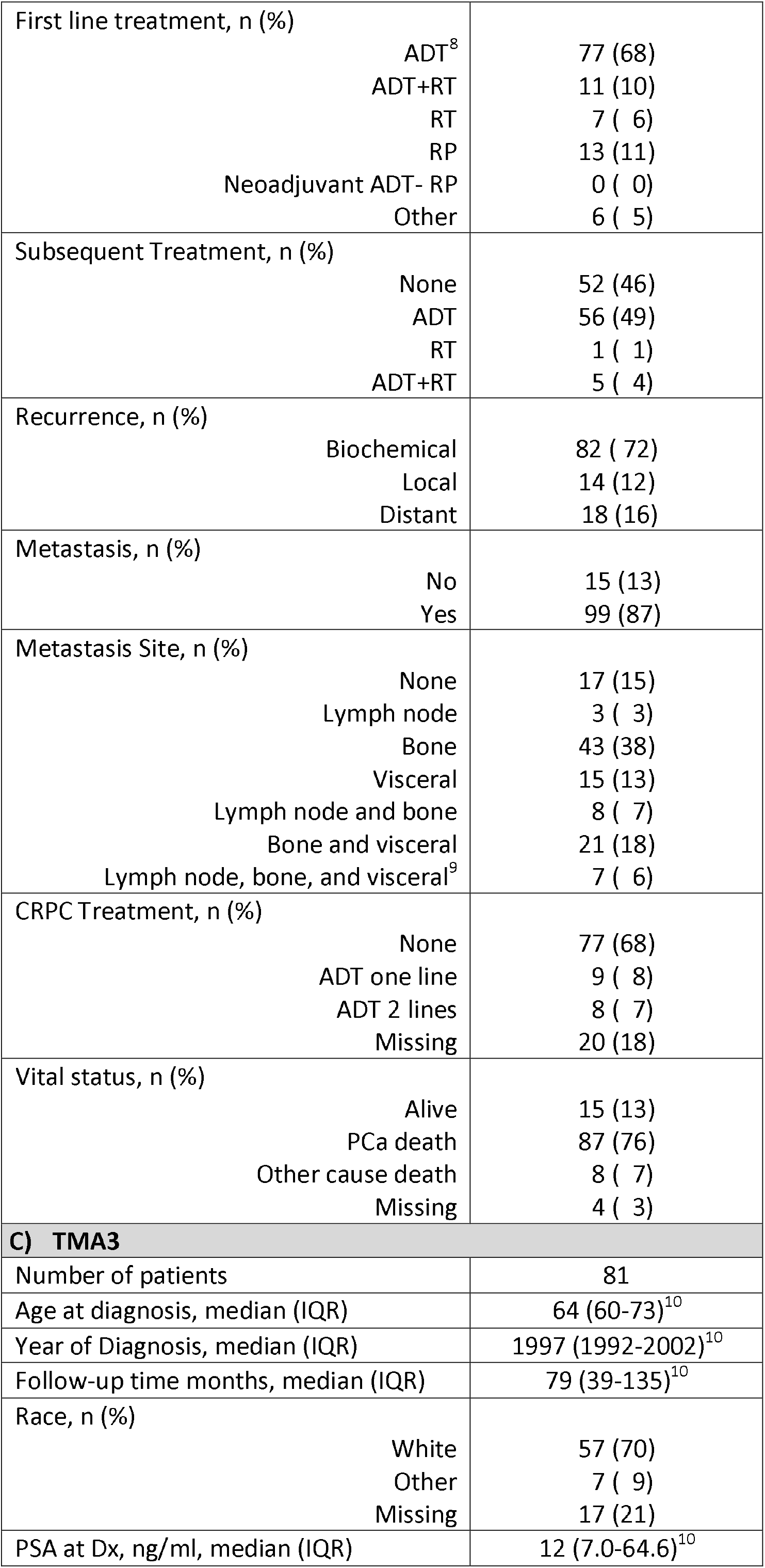
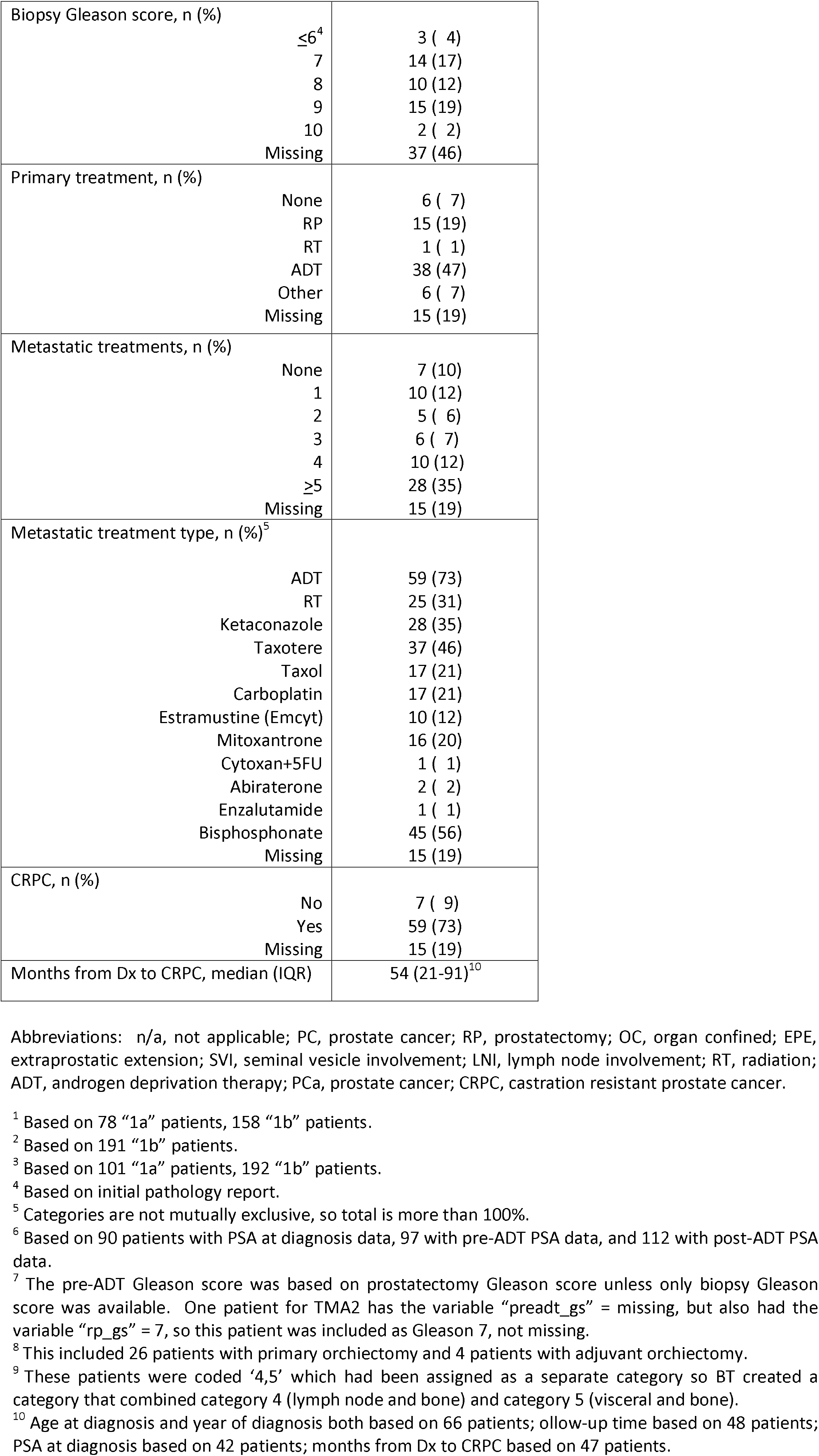
Demographic and clinical variables of the UTMA series

### Landmark PC markers

In the current study, as proof of concept, we used the test TMAs that will be provided, as a first assay control, to all researchers with approved TMA requests. These TMAs consist of a subset of cases included in the full TMAs. Test TMAs of each series were stained at a single institution for 6 IHC-based biomarkers. These markers were chosen because they are either selectively expressed in prostate cancer compared to most other cancer types (e.g. PSA, NKX3.1, PSMA, ERG, and AR), are known to be associated with disease progression (e.g. PTEN), or are a prostate cancer drug target (AR) (22–24). Furthermore, well-validated assays exist that can be performed using automated IHC staining, which are generally robust when considering variables relevant to the current study such as wide variability in tissue block age, tissue fixation extent, tissue handling, processing and storage (e.g. for PTEN, see(21)). **Figure 1** shows an example of IHC and H&E staining for each of the 6 IHC markers across Test TMA1. Representative images from Test TMAs 2 and 3 are shown in **Figures 2a and 2b**. All xenograft tissues stained as expected for each marker (**Supplemental Table 2**), providing excellent quality assurance for the IHC staining. For example, LNCaP and CWR22rv1 were positive for AR, NKX3.1 and PSMA and PC3 and DU145 were negative for these markers. All xenografts were negative for ERG, LNCaP and PC3 were negative for PTEN and DU145 and 22Rv1 were positive for PTEN.

**Figure 1.**
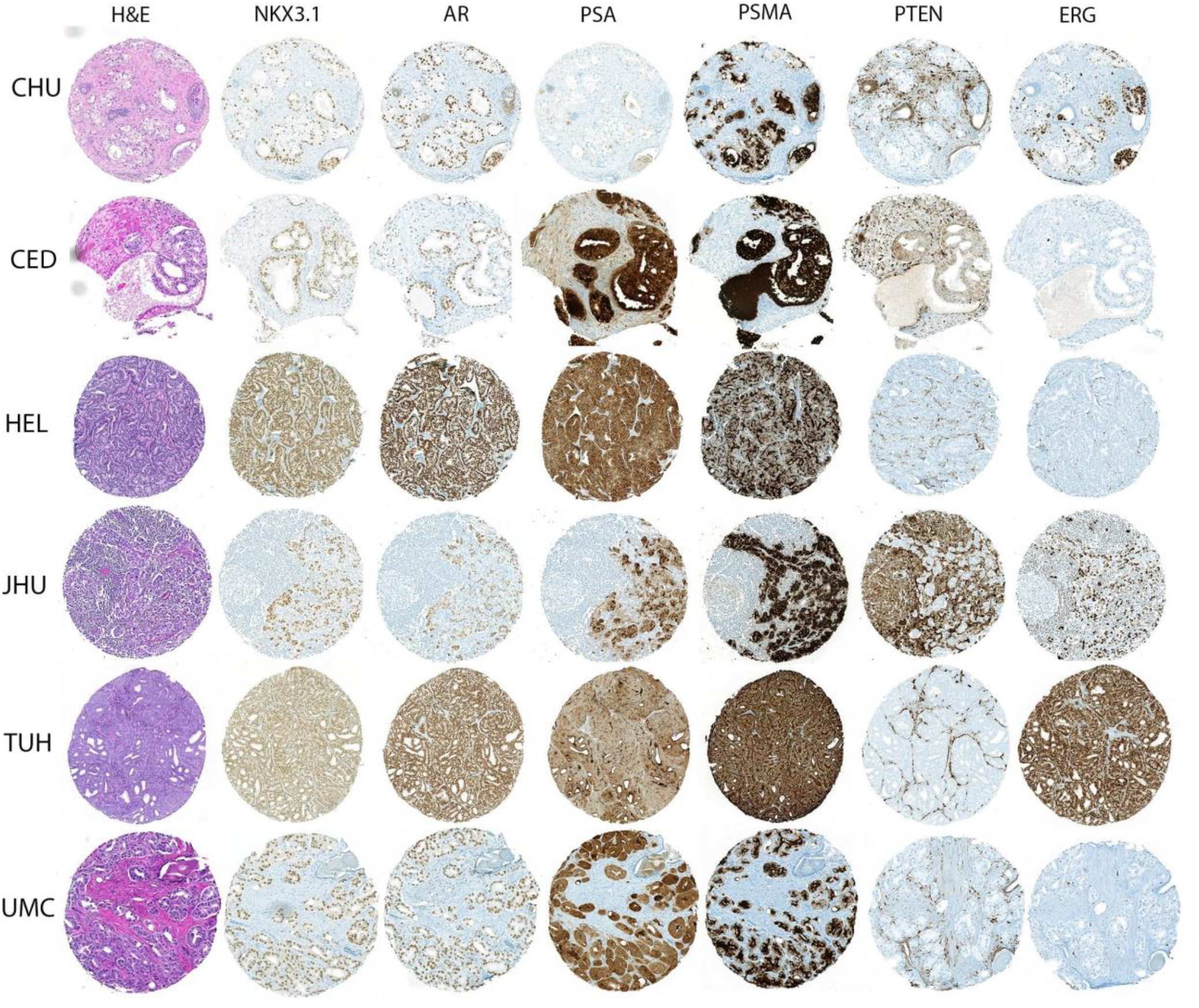
Representative low power view of staining of TMA cores from test-TMA 1. Each row shows staining from a single adjacent TMA core chosen from either a primary tumor or lymph node metastatic site from the indicated institutions.

**Figure 2.**
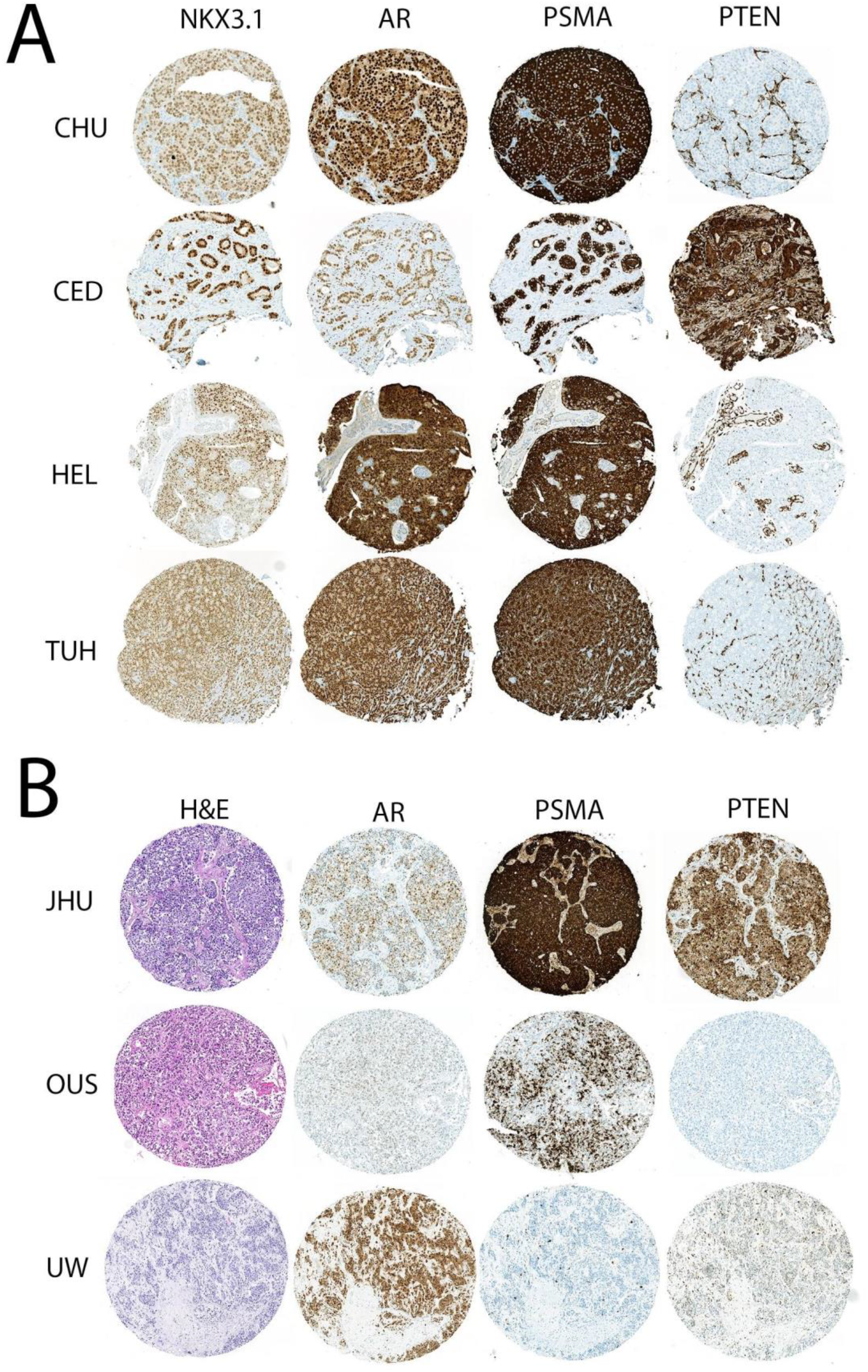
**(A)** Representative low power images from Test TMA 2. Each row shows staining from a single adjacent TMA core from the indicated institutions. **(B)** Representative images from Test TMA 3. Each row shows staining from a single adjacent TMA core from the indicated institutions.

**Table 3** shows the results for the fraction of TMA spots that were scored in each category as negative, positive-heterogeneous, positive-homogeneous (for ERG, AR, PSA, NKX3.1 and PSMA), or, for PTEN as positive, loss-heterogeneous or loss-homogeneous. For Test TMA1, using the primary tumors and metastatic lymph nodes only (e.g. without the xenografts included), as expected most tumors were positive for AR, PSA and NKX3.1 and PSMA (**Table 3**). Also, as seen in a number of prior studies, primary tumors or lymph node metastases that were ERG positive more commonly had PTEN loss (65.1% of cases with any ERG positive staining had any PTEN loss) than such tumors that were ERG negative cases (42.1% of cases with any ERG negative staining had any PTEN loss) (**Table 4**, P = 0.037, Chi2).

**Table 3.** Staining Results for Test-TMAs 1, 2 and 3.

| Biomarker | Test-TMA1 |  |  | Test-TMA2 |  | Test-TMA3<sup>1</sup> |  |  |
| --- | --- | --- | --- | --- | --- | --- | --- | --- |
|  | Tissue Type |  |  | Tissue Type |  | Tissue Type |  |  |
|  | Primary | LN Met | Xenograft | RP/TURP | Xenograft | Primary | Metastasis | Xenograft |
| PSMA Staining, n (%) |  |  |  |  |  |  |  |  |
| Negative | 0 (0) | 1 (3.1) | 11 (50) | 2 (4) | 7 (50.0) | 1 (12.5) | 24 (16) | 5 (41.7) |
| Positive heterogeneous | 19 (27.9) | 4 (12.5) | 4 (18.1) | 13 (26.0) | 2 (14.3) | 3 (37.5) | 24 (16) | 2 (16.7) |
| Positive homogeneous | 49 (72.1) | 27 (84.4) | 7 (31.8) | 35 (70) | 5 (40.0) | 4 (50) | 102 (68) | 5 (41.7) |
| **Total** | **68** | **32** | **22** | **50** | **14** | **8** | **150** | **12** |
| PTEN Staining, n (%) |  |  |  |  |  |  |  |  |
| Positive | 35 (54.2) | 13 (34.6) | 8(40) | 26 (48.2) | 8 (57.1) | 0 (0) | 14 (9.3) | 5 (35.7) |
| Loss heterogeneous | 2 (2.8) | 2 (7.7 ) | 1 (5) | 1 (1.9) | 1 (7.1) | 2 (25) | 7 ( 4.7) | 2 (14.4) |
| Loss homogeneous | 31 (43) | 15 (57.7) | 11(55) | 24 (44.4) | 5 (35.7) | 6 (75) | 127 (84.6) | 7 (50) |
| Cannot determine<sup>2</sup> | 0 (0) | 0 (0) | 0 (0) | 3 ( 5.6) | 0 (0) | 0 (0) | 2 (1.3) | 0 (0) |
| **Total** | **68** | **30** | **20** | **54** | **14** | **8** | **150** | **14** |
| ERG Staining, n (%) |  |  |  |  |  |  |  |  |
| Negative | 39 (54.9) | 22 (68.9) | 21 (95.2) | 29 (54.7) | 13 (100.0) | 7 (87.5) | 105 (70.0) | 12 (85.7) |
| Positive heterogeneous | 7 ( 9.9) | 5 (15.6) | 0 (0) | 9 (17.0) | 0 (0) | 1 (12.5) | 21 (13.9) | 2 (14.3) |
| Positive homogeneous | 25 (35.2) | 5 (0) | 0 (0) | 12 (22.6) | 1 (7.1) | 0 (0) | 19 (12.6) | 0 (0) |
| Cannot determine<sup>2</sup> | 0 (0) | 0 (0) | 0 (0) | 3 (5.7) | 0 (0) | 0 (0) | 6 (4) | 0 (0) |
| **Total** | **71** | **32** | **21** | **53** | **14** | **8** | **151** | **14** |
| AR Staining, n (%) |  |  |  |  |  |  |  |  |
| Negative | 0 (0) | 1 (3.1) | 10 (47.6) | 0 (0) | 6 (42.9) | 5 (62.5) | 24 (16) | 9 (64.3) |
| Positive heterogeneous | 13 (18.3) | 6 (18.8) | 3 (14.3) | 1 (1.9) | 2 (14.3) | 2 (25) | 19 (12.7) | 0 (0) |
| Positive homogeneous | 58 (81.7) | 25 (78.1) | 8 (38.1) | 51 (98.1) | 6 (42.9) | 1 (12.5) | 107 (71.3) | 5 (35.7) |
| **Total** | **71** | **32** | **21** | **52** | **14** | **8** | **150** | **14** |
| NKX3.1 Staining, n (%) |  |  |  |  |  |  |  |  |
| Negative | 2 ( 2.8) | 4 (12.5) | 10 (47.6) | 3 ( 5.6) | 6 (46.2) | 7 (87.5) | 75 (51.7.0) | 8 (61.5) |
| Positive heterogeneous | 21 (29.6) | 5( 15.6) | 7 (33.3) | 16 (29.6) | 6 (46.2) | 1 (12.5) | 35 (24.1) | 2 (15.3) |
| Positive homogeneous | 48 (67.6) | 23 (71.9) | 4 (19.1) | 35 (64.8) | 1 (7.7) | 0 (0) | 35 (24.1) | 3 (23.1) |
| **Total** | **71** | **32** | **21** | **54** | **13** | **8** | **145** | **13** |
| PSA Staining, n (%) |  |  |  |  |  |  |  |  |
| Negative | 1 ( 1.3) | 2 ( 6.3) | NA<sup>3</sup> | 8 (15.1) | NA<sup>3</sup> | 2 (25) | 48 (32.4) | NA<sup>3</sup> |
| Positive heterogeneous | 30 (41.7) | 5 (15.6) |  | 20 (37.7) |  | 2(25) | 42 (28.4) |  |
| Positive homogeneous | 41 (56.9) | 25 (78.1) |  | 23 (43.4) |  | 4 (50) | 58 (39.1) |  |
| **Total** | **72** | **32** |  | **51** |  | **8** | **148** |  |

<sup>1</sup>: There are 6 metastatic tissues each from 20 patients from UW, 4 each from 5 patients from JHU, and 2-4 metastatic sites from each of 4 patients from OUH and 2 samples of matched prostates from each of these patients from OUH.
<sup>2</sup> If there was a complete absence of staining in tumor cells and surrounding stromal cells, then that given TMA spot was considered inadequate for scoring and was not included.
<sup>3</sup> The anti-mouse antibody used for PSA is unable to be assessed on mouse xenografts.

**Table 4.** Correlation between PTEN loss and ERG status.

| Biomarker | ERG Status |  |  |  |
| --- | --- | --- | --- | --- |
|  | Negative | Positive<br>Heterogeneous | Positive<br>Homogeneous | P-value<sup>1</sup> |
| PTEN Status, n (%) |  |  |  |  |
| Loss heterogeneous | 4 (7.0) | 0 (0.0) | 0 (0) | **0.032** |
| Loss homogeneous | 20 (35.0) | 9 (75.0) | 17 (59.0) | - |
| Positive homogeneous | 33 (58.0) | 3 (25.0) | 12 (41.0) | - |
| Total | 57 | 12 | 29 | - |

<sup>1</sup> Chi-squared test.

**Table 3** also shows the staining and scoring results for Test TMA2, which consisted of tissue samples from RP, TURP as well as the control tissues and xenografts. Excluding the xenografts, most cases of carcinomas stained adequately, and most were positive for the 4 prostate enriched biomarkers (AR, PSA, NKX3.1 and PSMA). ERG was positive in at least a fraction of tumor cells in 42 % of cases and PTEN was lost homogeneously in 47% of cases and lost heterogeneously in 1 additional case. PTEN loss was again more common in ERG positive cases (55% of cases with any ERG positive staining had any PTEN loss versus 44.8% of ERG negative cases had any PTEN loss), although this was not significant (P=0.48, Chi2).

Test TMA3 consists of metastatic tumor tissues from autopsies from three institutions as well as a small number of primary prostate carcinomas from one of the sites and the control xenograft tissues. Focusing on the metastatic autopsy tumor spots, the majority (84%) had at least some positive staining for PSMA (84%), AR (84%), and PSA (68%). NKX3.1 was positive in 48.3% of metastatic TMA cores. This relatively low fraction of cases staining positively using the NKX3.1 antibody may relate to either less frequent presence of NKX3.1 protein in these very late stage disease tissues, or, poor preservation of its protein target as a result of prolonged warm ischemic time. Deciphering which of these alternatives may be correct is difficult and represents an inherent limitation of using autopsy tissue for some biomarkers (see discussion). ERG was positive in 28% and PTEN was lost in 90.5% of metastatic cores in test TMA3. The majority of metastatic TMA cores with PTEN loss showed homogeneous loss (86%). This rate of PTEN loss is remarkably high and it will be interesting to determine if this rate of loss holds in the full GAP1 TMAs when PTEN is evaluated in those. Unlike in test TMAs 1 and 2, the fraction of ERG positive cases with PTEN loss was lower than in ERG negative cases in test TMA3 (P < 0.0001).

### Resource available for PC community - Process

The process to access the UTMA resource is outlined in **Supplemental Figure 1**. As part of a collaboration, the biomarker proposal form will be located on the Prostate Cancer Biorepository Network (PCBN) (25) website (www.prostatebiorepository.org). Since the ultimate objective of the UTMA GAP1 program is to identify biomarkers and combinations thereof that would be helpful for clinicians to refine patient management, end users are required to agree to return their raw results (scoring results and protocols) back to the GAP1 coordinating center either after publication, or after 6 months from scoring completion, whichever is sooner.

## Discussion

Tissue microarrays represent an efficient format to profile biomarkers from many different tumors at the same time. However most studies use either a small number of samples or only include cases from a single institution. With the vision of building a unique resource for prostate cancer researchers, Movember brought together 19 principal investigators from 13 institutions based in different continents. Through the development of specific guidelines, model documents for TMA design and clinical data collection using an harmonized dictionary, we successfully created a unified resource representing different disease states of prostate cancer.

Demographics of the TMA series showed that the cohorts are representative of prostate cancer patients in terms of age at diagnosis. The majority of the patients in the cohort are white. Therefore, the generalizability of these TMA resources are limited somewhat in terms of race. The serum PSA level at diagnosis was generally lower than expected for TMA1, given that only 10% of patients within the PSA range of TMA1 have LNI based on commonly used predictive tables (26). However, this may be partially explained by half of the patients having only one or two positive LN. The other clinico-pathological variables such as Gleason score, staging and pathological findings are more in line with what would be expected for this cohort. TMA2 and TMA3 cohorts are composed of patients diagnosed with PC more than two decades ago and clinico-pathological data are representative of the type of disease included in this series. In addition, treatment provided reflects the reality of limited therapeutic options for patients with advanced/castration resistant prostate cancer during this period.

Although these results are preliminary because of small sample sizes in the test TMAs, biomarker expression patterns and frequencies were mostly as expected based on prior work. While some of the findings were somewhat unexpected, such as the very high rate of PTEN loss in the autopsy samples (86%), current studies by our group are ongoing in which we are examining these markers in the full TMAs and details regarding them will be published separately. In concurrent preliminary analyses of PTEN and ERG, we found that any PTEN loss was more common in cases with ERG expression in Test TMAs 1 and 2 as has been seen previously (8), but present at an even rate between ERG positive and ERG negative specimens in the CRPC autopsy samples in Test TMA 3. Taken together, these results indicate that the staining for the biomarkers employed were robust in various types of tissues processed separately at multiple institutions and that the staining results are consistent with previous reports.

Some limitations of our study should be noted. The resource is composed entirely of samples punched from older FFPE specimens, in which there tends to be decreases in RNA-ISH signals over time (18) and hence this, as with many other TMAs, may not be highly suitable for *in situ* hybridization studies for RNA in general. Furthermore, a common concern of the use of TMA’s is whether a small sampled area can accurately capture heterogeneity in the wider specimen. While this issue is always a factor in all biomarker studies of tissues, including using biopsy specimens, this was accounted for to a certain extent in TMA design by selecting multiple TMA cores per specimen. Furthermore, a number of prior studies have addressed this issue and have generally reported that TMAs are often quite robust to tissue heterogeneity, especially when multiple replicates are included (27–29). Finally, rapid autopsy tissue has limitations. One limitation is that the time between death and tissue fixation is variable and can be from several hours up to 24 hours in this study. While some biomarkers may be retained fully with such post-mortem intervals in which vascular tissues were studied (30), it cannot be known which others may not be robustly retained without experimental data on that given biomarker. In addition, bone lesions at autopsy underwent decalcification, generally in formic acid. While prior studies by one of our groups have not identified negative effects of decalcification on expression of the analytes in prior studies using similar samples (31–35), analytes that we have not studied, such as phosphoproteins, could potentially be altered. Another limitation of these specimens is that they were collected from patients with CRPC no later than 2013, before late generation androgen deprivation therapy was widely used. Consequently, only two of the fifty patients received modern anti-androgens (abiraterone or enzalutamide) for a significant time during their treatment.

In summary, we have developed a set of TMAs from a multi-institutional initiative supported by the Movember Foundation. This effort focused on rare but clinically important samples made possible through an international collaboration. It required leveraging existing institutional resources and multidisciplinary prostate cancer expertise across multiple continents to provide a unique resource that will serve the wider prostate cancer research community to support discovery based research and enhance the overall impact of biomarker validation studies.

## Data Availability

The process to access the UTMA resource is outlined in Supplemental Figure 1. As part of a collaboration, the biomarker proposal form will be located on the Prostate Cancer Biorepository Network (PCBN) (25) website (www.prostatebiorepository.org).

## Acknowledgements

We thank all patients for providing specimens and data to the UTMA-affiliated biobanks. We thank ÉloÏse Adam-Granger, Kathy Doan, Andrée-Anne Grosset, Belinda Nghiem, Olov Øgren, Tiina Vesterinen, RRCancer-CRCHUM prostate cancer biobank staff and the CRCHUM molecular pathology platform for TMA preparation, sample collection and clinical data integration and Yasser Amhdak, Lauri Elo, Lori Kollath for sample and clinical data collection. We thank the rapid Autopsy Team, Celestia Higano, Pete Nelson, Bruce Montgomery. Evan Yu, Elahe Mostaghel, Paul Lange, Robert Vessella, Xiaotun Zhang, Martine Roudier for contributing to the rapid autopsy programs and Gayle Walters for legal support.

## Notes

**Financial support:** The creation of this resource was funded by Movember. JHU is supported by U.S. NIH/NCI SPORE in Prostate Cancer: P50CA58236, the U.S. Department of Defense Prostate Cancer Research Program (PCRP): W81XWH-18-2-0015 to A.M.D. The Johns Hopkins Sidney Kimmel Comprehensive Cancer Center Oncology Tissue Services Laboratory supported by U.S. NIH/NCI P30 CA006973. CRCHUM is supported by the FRQS. Biobanking at the CRCHUM was done in collaboration with the Réseau de recherche sur le cancer of the Fonds de Recherche Québec - Santé (FRQ-S) affiliated to the Canadian Tissue Repository Network (CTRNet). F.S. the Raymond Garneau Chair in Prostate Cancer. D.T. is supported by the Chercheure boursière clinicienne niveau junior 2 of the FRQ-S awards. UW tissue acquisition was supported by U.S. NIH/NCI SPORE in Prostate Cancer: P50CA97186 and the UW Institute for Prostate Cancer Research.

### Competing Interest Statement

The authors have declared no competing interest.

### Author Declarations

All centers received ethical review board permission to use patient material and conduct this study Comite d ethique de la recherche du CHUM (CE.14.128) the Institutional Review Board of the : JHU School of Medicine s the UMBMC the VA Greater Los Angeles (Department of Veterans Affairs PCC#2015-040408) the Ethics Committee of Hospital District of Helsinki and Uusimaa (84/13/03/00/2014; 3 30.01.2015) the Hospital District of Southwest Finland (number T206/2014) National Supervisory Authority for Welfare and Health for HUS and TYKS (VALVIRA 8008/06.01.03/2014) the Regional Committees for Medical and Health Research Ethics for OUH (REC 2013/1713) and Center for Healthcare Ethics for Cedar Sinai (Pro00033387 and Pro00020577). Patient informed consents were obtained as required by individual institutional ethical review boards (CHUM CE12.216 HUS TYKS and University of Washington Cedar Sinai) or waivers were granted (JHU OUH both VA Health Care System and UMBMC).

