## Supplementary figures and images for "The Movember Global Action Plan 1 (GAP1) - Unique Prostate Cancer Tissue Microarray Resource"

### Supplementary Figure 1

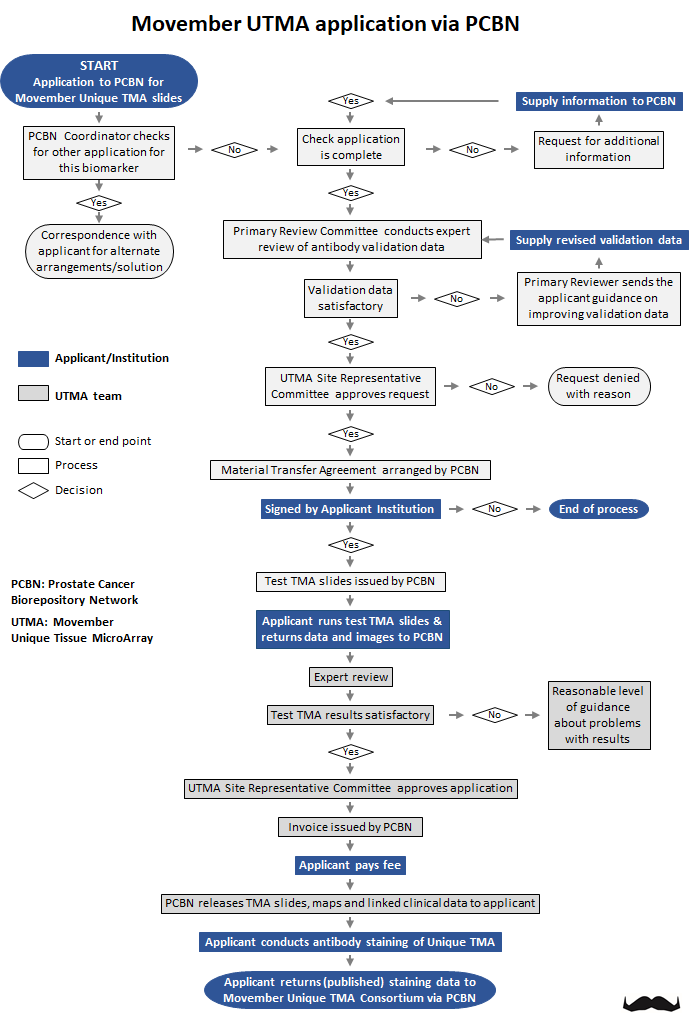

**Supplemental Figure 1.** UTMA resource access procedure.
