## Supplementary Table 2 for "The Movember Global Action Plan 1 (GAP1) - Unique Prostate Cancer Tissue Microarray Resource"

**Supplemental Table 2. Xenograft Staining Results in Test TMAs.**

|  | **Xenograft** | | | |
| --- | --- | --- | --- | --- |
| **IHC Marker** | **LNCaP** | **PC3** | **22Rv1** | **DU145** |
| AR | + | - | + | - |
| NKX3.1 | + | - | + | - |
| PSMA | + | - | + | - |
| PTEN | - | - | + | + |
| ERG | - | - | - | - |

- Designates positive staining in tumor cells. - designates negative staining in tumor cells. The anti-mouse antibody used for PSA is unable to be assessed on mouse xenografts.
